# Epidemiological characteristics and incubation period of SARS-CoV-2 during the 2020-2021 winter pandemic wave in north China: an observational study

**DOI:** 10.1101/2021.05.08.21256896

**Authors:** Tiantian Liu, Zijian Chen, Jin Xu

## Abstract

As the emergence of new variants of SARS-CoV-2 persists across the world, it is of importance to understand the distributional behavior of incubation period of the variants for both medical research and public health policy-making. We collected the published individual level data of 941 patients of the 2020-2021 winter pandemic wave in Hebei province, north China. We computed some epidemiological characteristics of the wave and estimated the distribution of the incubation period. We further assessed the covariate effects of sex, age and living with a case with respect to incubation period by a model. The infection-fatality rate was only 0.1%. The estimated median incubation period was at least 22 days, significantly extended from the estimates (ranging from 4 to 8.5 days) of the previous wave in mainland China and those ever reported elsewhere around the world. The proportion of asymptomatic patients was 90.6%. No significant covariate effect was found. The distribution of incubation period of the new variants showed a clear extension from their early generations.

## 1. Introduction

Since the outbreak of COVID-19 pandemic in December 2019 from Wuhan China, there have been more than 12 million confirmed cases and 2 million deaths reported all over the world.^1^ The SARS-CoV-2 appeared to evolve in various ways to survive in human as mutations had been found such as the British mutation,^2^ the Brazil variant,^3^ and the South Africa variant,^4^ among others.

The incubation period (IP), defined as the time between infection and symptom onset, serves as a key epidemiological parameter for medical researchers to understand the viral virulence and for health authorities to make appropriate policies. There have been intensive studies of incubation period of SARS-CoV-2 for the first pandemic wave in China with reported median of IP ranging from 4 to 8.5 days^5–19^ and possible covariate effects such as age,^10,19^ sex^19,20^ and travel history.^18^ The current general policy for the quarantine time by WHO and many governments is 14 days.

By late April 2020, mainland China had almost eradicated the domestic pandemic with about 90 consecutive days of domestic cases under 10 per day.^21^ From 11^th^ June 2020, there were several isolated clusters in small scale (with maximum domestic cases of 257) such as in Beijing, Qingdao (in Shandong province), Dalian (in Liaoning province), Suifenhe (in Heilongjiang province), until the 2020-2021 winter pandemic wave took place in Hebei province in north China. This pandemic wave caused lockdown of three cities (Shijiazhuang, Xingtai, Langfang) of population more than 20 million, quarantine of more than 20 thousand people, and one death.^22^ All these cases were considered to be imported cases with variants of SARS-CoV-2 from Europe or Russia after generations from the original type in December 2019.^23^

In this article, we conducted an observational study based on the published individual level data of 941 patients to study the distributional behavior of incubation period of the new variants and some epidemiological characteristics of interest. Our study aims to (i) shed lights on the evolution of the SARS-CoV-2 from the statistical perspective of the distribution of incubation period and (ii) provide bases for public health policy-making.

## 2. Methods

### 2.1 Study design and data

This study is classified as an observational study or public health surveillance and is exemplified from ethical review or informed consent by the institutional review board.

Since 3^rd^ January 2020, when the first case of SARS-CoV-2 was diagnosed in Shijiazhuang of Hebei province, the national health commission of Hebei province started to publish individual patient narratives regarding the transmission timeline and the tracing information along with some important covariates such as sex, age, and the status of living with a case, etc. We collected such data from 941 patients of Hebei province in north China till the last publication on 4^th^ February 2021. The summary of the data sources is provided in Appendix A.

We extracted relevant data from the patient narratives with great care. Specifically, let T_0_ denote the date of infection, e.g., a date who attended a wedding which turned out to be a transmission occasion. When T_0_ could not be precisely recalled, let [W_0_, W_1_] denote the time window where T_0_ belongs, which is either recalled by patient or derived from known information. For example, W_0_ is the date when one returns from another city and W_1_ is the date when one gets self-isolated or quarantined. Let T_1_ denote the date of symptom onset, where symptom includes fever, dry cough, tiredness, etc., confirmed by physicians. So IP is simply T_1_–T_0_. At last, let T_2_ denote the date of being diagnosed by nucleic acid test. Note T_2_ is always observed.

In the later period of this wave, due to mass quarantine, 249 of 941 patients did not have information about the date of infection. I.e., neither T_0_ nor the window [W_0_, W_1_] were available. Therefore, only 690 subjects were used in analyzing the distribution of IP. According to the actual observations of T_0_ and T_1_, we categorized the data into the following four types. Type I: Both T_0_ and T_1_ are observed. And T_2_ is greater than T_1_. Type II: T_0_ is observed. But T_1_ is censored by T_2_, i.e., one is diagnosed without symptom. Type III: T_1_ is observed. But T_0_ is left truncated by interval [W_0_, W_1_]. Type IV: Neither T_0_ and T_1_ are observed, i.e., T_0_ is left truncated by [W_0_, W_1_] and right censored by T_2_ simultaneously. We illustrate the four types of data in Figure 1 and provided examples for each type in Appendix B. At last, we defined the status of living with a case (yes/no, coded as 1/0), where the confirmed case can be the spouse or a relative such as grandma, brother, etc.

**Figure I:**
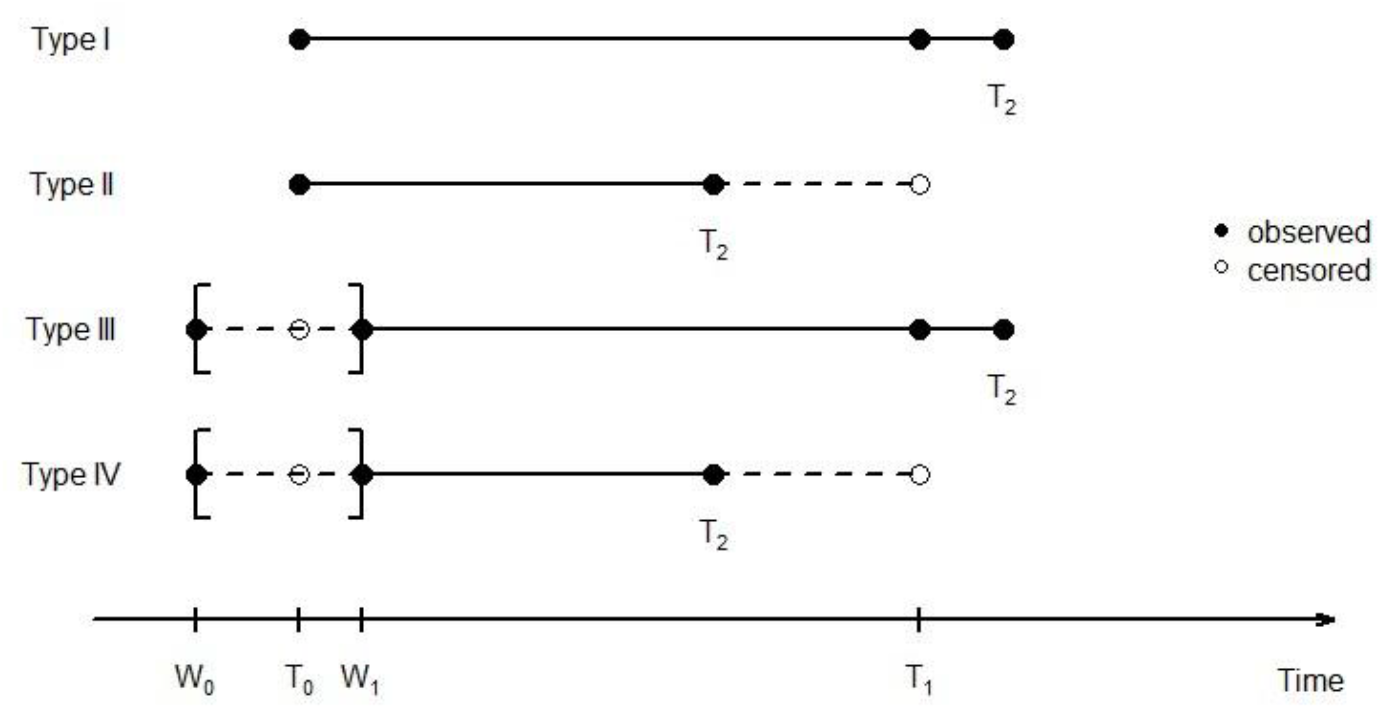
Illustration of the four types of observations of T_0_ and T_1_ with associated [W_0_, W_1_] (if applicable) and T_2_, where the solid bullet stands for an actual observation and the empty circle stands for censoring

### 2.2 Statistical model and analysis

We treated the incubation period as time-to-event (survival) random variable subject to right censoring. Here event stands for symptom onset and right censoring occurs when a patient is diagnosed without symptoms. The combined type I and type II data constituted the main body of such data. For type III data with left truncation, we treated them as type II data in a reverse direction, i.e., treat IP was censored from left. For type IV data, we approximated T_0_ by W_1_ so that to (under)estimate IP in a conservative way as Nie^14^. This is in contrast to the liberal estimation by replacing T_0_ by W_0_9 and the robust estimation by replacing T_0_ by the middle point of W_0_ and W_1_.^19^

Unlike many statistical analysis methods to estimate the distribution of IP using parametric models such as lognormal, Weibull and exponential models,^5,12,24^ we used the traditional non-parametric Kaplan-Meier method to estimate the probability of being asymptomatic (survival function) and subsequently the median IP.^25^ We provided a sensitivity analysis using parametric models to examine the impact of modeling.

We used the well known semi-parametric Cox model to assess the significance of covariate effect.^25^ All tests were two sided with significance level 0.05. All statistical analyses were carried out by statistical software R (version 4.0.4) through the package ‘Survival’ primarily.

## 3. Results

### 3.1 Summary of some epidemiological characteristic

A total of 941 cases from 2^nd^ January 2021 to 3^rd^ February 2021 (throughout the pandemic wave of 32 days) in Hebei province were collected as described in Methods section. Figure II shows the numbers of daily diagnosed cases along with the associated reproduction number R_0_. It is seen that the pandemic wave climbed exponentially in the first 10 days and plateaued at about 90 cases per day for four days before a rapid drop. The overall R_0_ for the wave was around 0.84–1.0, depending on the estimation methods.^26^ The quick control of the pandemic was largely due to the prompt measures of mass test and lockdown.

**Figure II:**
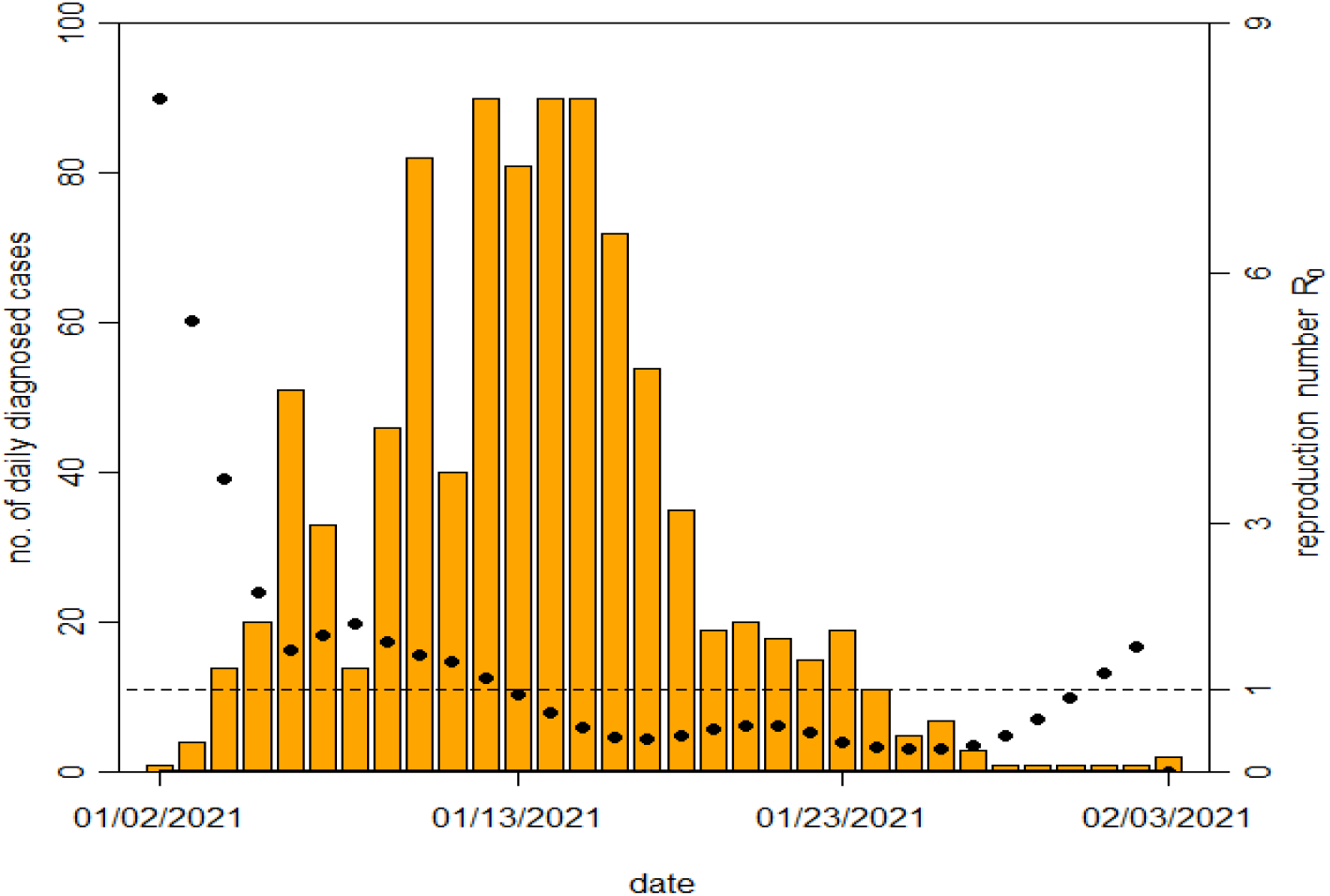
Number of daily diagnosed cases (left axis) and reproduction number R_0_ (right axis) of the studying pandemic wave

Table I summarizes some epidemiological characteristics of the data. The findings were as follows. (i) Individuals of age 31-60 were more likely to be infected compared with other age groups. Similar age-based difference was detected by Zhang.^26^ (ii) Women had a larger infection rate (58.7%, 552 of 941) in this wave than that (41.7%, 389 of 941) of men. (iii) 19.3% (182 of 941) of the patients were living with a case. (iv) The proportion of asymptomatic patients was as high as 90.6% (853 of 941), nearly ten times of that (9.4%, 88 of 941) of the symptomatic patients. While, this proportion of asymptomatic patients in the previous wave (as of 11th February 2020) was just 1.2%.^28^ (v) The median time period between symptom onset and being diagnosed was 2 days. Note that results in (iv) and (v) were largely due to the mass test. (vi) 59% (573 of 941) of the patients had at least one negative outcome of nucleic acid test before being diagnosed lately. 20.2% (190 of 941) of them even had experienced more than three times of such “false” negative results. This remarkable proportion of “false negative” corroborated with the finding in (iv) and could be attributed to the sensitivity of the test kit and more likely to the prolonged incubation period when the variants of the virus was not detectable.^29^

**Table I:** Epidemiological characteristics of the study patients.

| Characteristic | Statistic |
| --- | --- |
| N | 941 |
| Age |  |
| Median (IQR) - year | 46 (30) |
| Minimum - year | 0.5 (6 months) |
| Maximum - year | 91 |
| Distribution - no. (%) |  |
| 0 – 10 | 77 (8.2) |
| 11 – 20 | 78 (8.3) |
| 21 – 30 | 97 (10.3) |
| 31 – 40 | 164 (17.4) |
| 41 – 50 | 129 (13.7) |
| 51 – 60 | 177 (18.8) |
| ≥ 60 | 219 (23.3) |
| Sex - no. (%) |  |
| Male | 389 (41.3) |
| Female | 552 (58.7) |
| Exposure status - no. (%) |  |
| Living with a case | 182 (19.3) |
| Others | 759 (80.7) |
| Symptom status before being diagnosed - no. (%) |  |
| Symptomatic | 88 (9.4) |
| Asymptomatic | 853 (90.6) |
| Days between symptom onset and being diagnosed |  |
| Minimum | 0 |
| Maximum | 11 |
| Mean | 2.4 |
| Median | 2 |
| Number of negative tests before being diagnosed - no. (%) |  |
| 0 | 386 (41.0) |
| 1 | 64 (6.8) |
| 2 | 146 (15.5) |
| 3 | 155 (16.5) |
| 4 | 79 (8.4) |
| 5 | 48 (5.1) |
| 6 | 19 (2.0) |
| 7 | 21 (2.2) |
| 8 | 16 (1.7) |
| 9 – 12 | 7 (0.7) |

Among all 690 patients with at least one observation of T_0_ or T_1_, there were 27, 180, 61 and 422 patients of types I to IV, respectively. It is noted that in the earlier period of this wave (3^rd^ January, 2021 to 8^th^ January, 2021) most cases (122 out of 137) were reported in rural areas where the patients (many were local farmers) sought first medical aid in local clinics when the initial symptoms appeared. Some of them could not recall precisely when the infection contact was made and we had to estimate it by a broader time window. These patients became the major sources for types I and III data. As the pandemic spread into the cities, lockdown was imposed and mass tests were given. We found most asymptomatic cases of types II and IV data in the later time period.

### 3.2 Estimation of the distribution of incubation period and the median

We estimated the probability of being asymptomatic based on different aggregated types of data (Figure III), from which the estimated median IP were found to be greater than 22 days which was the lower limit of the 95% confidence interval. The actual median could be much larger as seen by Figure 3. The minimum observed incubation period was 3 days.

**Figure III:**
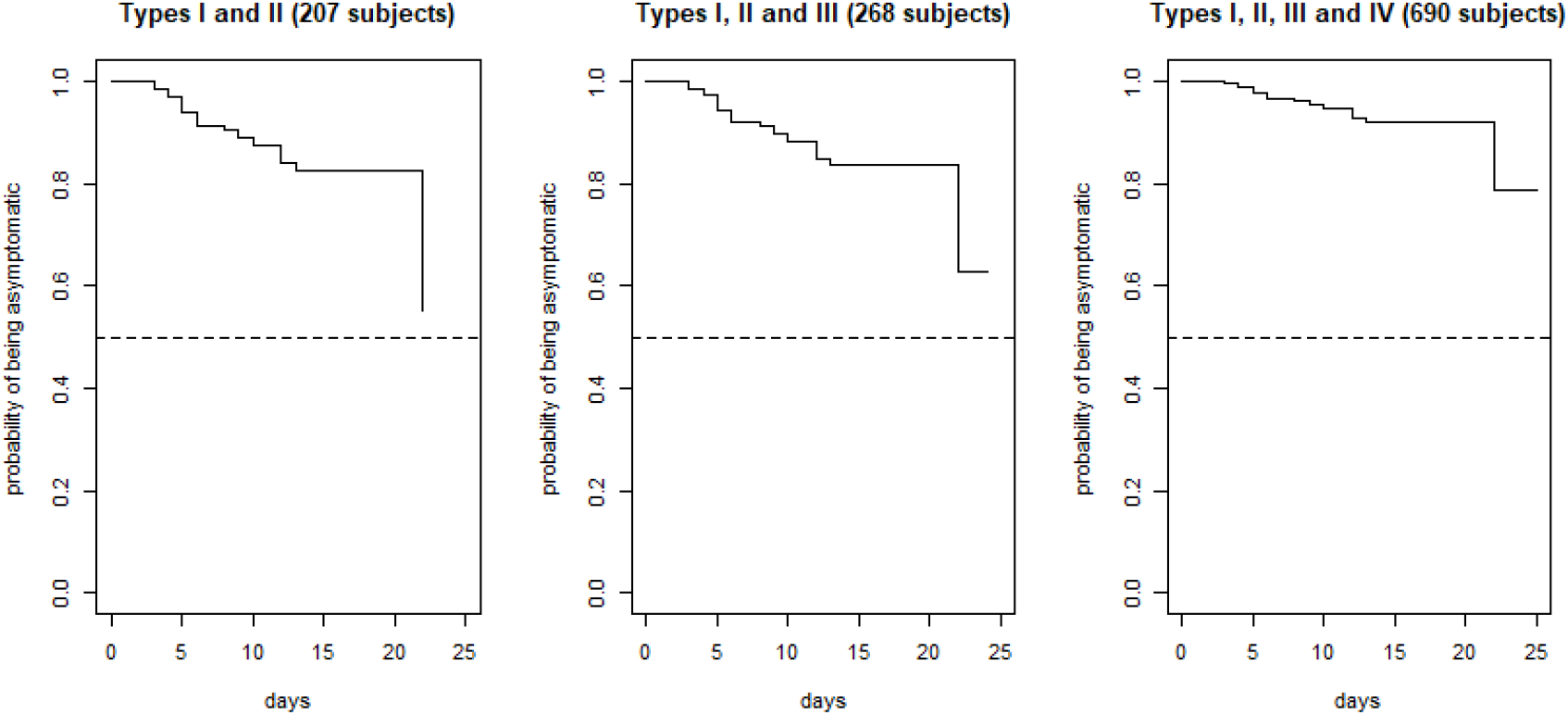
Kaplan-Meier estimates of probability of being asymptomatic based different aggregated types of data, where the horizontal dotted line at 0.5 does not cross with any estimated survival curves

We compared the estimate of the median IP to 13 reported counterparts over different time periods in Figure IV. The summary of the comparing estimates was provided in Appendix C. First, we noticed that there was a time gap of about eight months between this wave and the previous wave. Second, the median IP of this wave was significantly prolonged comparing to those in the first wave before May 2020. A meta-analysis based on seven out of 13 articles with information of confidence intervals showed the median IP of the previous wave was 5.6 days (95% CI 5.1–8.3). (See Appendix D for the detail.) This indicated possible virological changes of the variants of SARS-CoV-2 that evolved to adapt to the new biological environment.

**Figure IV:**
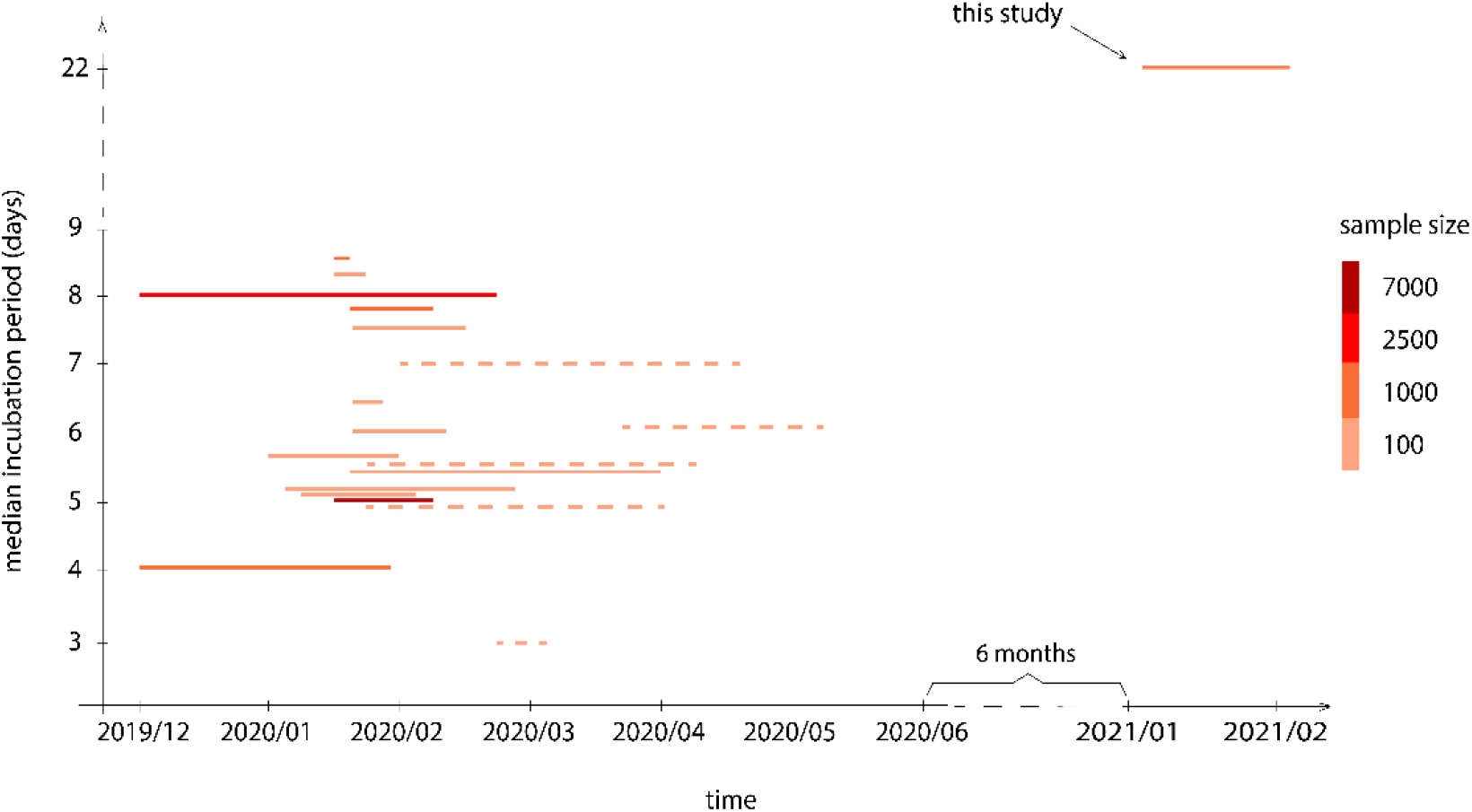
Comparison of the estimate of the median IP of the study wave and 13 reported estimates of median IP of the previous pandemic wave in China over different time periods (solid lines). Five estimates of the median IP based on the pandemics in India, Vietnam, South Korea, Singapore and Argentina were also included (dotted lines).

### 3.3 Covariate analysis

The covariate analysis by the Cox model found no significant covariate effect (with respect to IP) by sex, age or living with a case based on different aggregated data. It suggested that the IP of the new variants of SARS-CoV-2 exhibited indiscriminately with respect to the considered covariates. We also split the data into two age groups, namely “less than 60” and “no less than 60”. The log-rank test^25^ again found no difference of the distribution of IP between the two groups. (See all details in Appendix E.)

### 3.4 Sensitivity analysis

We used two commonly used parametric models, namely Weibull and lognormal models, to estimate the distribution of IP and the median. It was found that median IP was at least 29 days (95% CI 20.1– 42.3) under Weibull model and at least 37.6 days (95% CI 23.0–61.3) under lognormal model (Table II).

**Table II:** Estimates of the median IP and the corresponding 95% CI based on different aggregated types of data under Weibull and lognormal models.

| Aggregated types | Model | Median | 95% CI |
| --- | --- | --- | --- |
| I+ II | Weibull | 29.1 | [20.1, 42.3] |
|  | Lognormal | 37.6 | [23.0, 61.3] |
| I + II + III | Weibull | 30.2 | [20.8, 43.9] |
|  | Lognormal | 39.6 | [24.1, 65.0] |
| I + II + III + IV | Weibull | Infinity | Infinity |
|  | Lognormal | 71.5 | [38.0, 134.6] |

## 4. Discussion

Understanding the statistical distribution of incubation period is critical for studying the pathogenic process, dynamics of transmission, case fatality, and epidemic size. The derived parameters such as median incubation period and 95% percentile are key for public health policy decision such as isolating infected cases and quarantine period.

In this study, we presented a rather comprehensive analysis of the distributional characteristics of IP based on published data of the 2020-2021 winter pandemic wave in north China, where the variants were thought imported from Europe or Russia.

We found that the median IP was greater than 22 days. It was significantly extended from 5.6 days of the previous wave from December 2019 to April 2020. This large leap was reflected by the recent finding of He^30^ that 6.9% of population in Wuhan China was asymptomatic throughout the whole period (from infection to self-recovery). It is worth noting that the infection-fatality rate of this wave was only 0.1% (1/941). It might be attributed to the changed virulence of the variants and the timely treatment in early stage. This finding is corroborated by recent reports of imported cases, many of whom were diagnosed positive after two to three weeks of quarantine without any symptom. Our results showed no significant covariate effect of sex, age and living with a case. In particular, we found no significant difference in distribution of IP between groups of patients older than 60 and younger than 60.

The strengths of this study are twofold. First, our data set covered a complete pandemic wave of 941 cases and contained accurate patient level information. Second, by taking advantage of the large sample size, we adopted the non-parametric method to estimate the distribution of IP with high precision. In contrast to the parametric models, our approach is model free and therefore robust to the presence of extreme observations.

Although we made great effort in ascertaining the dates of infection and symptom onset, there inevitably existed biases due to recall and deduction from tracing information. Our approach here for estimation was conservative such as to underestimate IP by using W_1_ to approximate T_0_ when analyzing the type IV data.

In conclusion, the distribution of IP of the new variants of SARS-CoV-2 tends to have a heavy tail towards right with a prolonged median of at least 22 days. An extended period of three weeks by a combination of quarantine and self-isolation is recommended for asymptomatic individual from infected regions. Long term monitoring system is warranted to study the evolution of COVID-19.

## Data Availability

The source of data is provided in Appendix. The data collected for the study will be made available up on request.

## Author contributions

JX conceived and designed the study, oversaw the collection of the data, took responsibility for the accuracy of the analysis. All authors had full access to all of the data in the study. TL and ZC collected and analyzed the data. TL, ZC and JX wrote the first draft of the manuscript. All authors critically reviewed and revised the manuscript.

## Acknowledgments

This study was supported by Key Laboratory of Advanced Theory and Application in Statistics and Data Science – Ministry of Education, China.

