## Supplemental file for "Epidemiological characteristics and incubation period of SARS-CoV-2 during the 2020-2021 winter pandemic wave in north China: an observational study"

### A. List of data source of individual patient narratives

| Date (YYYY/MM/DD) | Province | City | N | Source webpage |
|---|---|---|---|---|
| 2021/01/02 | Hebei | Shijiazhuang | 1 | https://news.sina.com.cn/c/2021-01-03/doc-iiznezxt0358165.shtml |
| 2021/01/03 | Hebei | Shijiazhuang | 2 | https://news.sina.com.cn/c/2021-01-04/doc-iiznctkf0094732.shtml |
| 2021/01/03 | Hebei | Xingtai | 2 | https://new.qq.com/omn/20210104/20210104A02GQ700.html |
| 2021/01/04 | Hebei | Shijiazhuang | 11 | wsjkw.hebei.gov.cn/syyctplj/375192.jhtml |
| 2021/01/04 | Hebei | Xingtai | 3 | wsjkw.hebei.gov.cn/syyctplj/375191.jhtml |
| 2021/01/05 | Hebei | Shijiazhuang | 19 | wsjkw.hebei.gov.cn/syyctplj/375221.jhtml |
| 2021/01/05 | Hebei | Xingtai | 1 | wsjkw.hebei.gov.cn/syyctplj/375219.jhtml |
| 2021/01/06 | Hebei | Shijiazhuang | 50 | wsjkw.hebei.gov.cn/syyctplj/375261.jhtml |
| 2021/01/06 | Hebei | Xingtai | 1 | wsjkw.hebei.gov.cn/syyctplj/375260.jhtml |
| 2021/01/07 | Hebei | Shijiazhuang | 31 | wsjkw.hebei.gov.cn/syyctplj/375294.jhtml |
| 2021/01/07 | Hebei | Xingtai | 2 | wsjkw.hebei.gov.cn/syyctplj/375293.jhtml |
| 2021/01/08 | Hebei | Shijiazhuang | 14 | wsjkw.hebei.gov.cn/syyctplj/375333.jhtml |
| 2021/01/09 | Hebei | Shijiazhuang | 44 | wsjkw.hebei.gov.cn/syyctplj/375370.jhtml |
| 2021/01/09 | Hebei | Xingtai | 2 | wsjkw.hebei.gov.cn/syyctplj/375369.jhtml |
| 2021/01/10 | Hebei | Shijiazhuang | 77 | wsjkw.hebei.gov.cn/syyctplj/375391.jhtml |
| 2021/01/10 | Hebei | Xingtai | 5 | wsjkw.hebei.gov.cn/syyctplj/375390.jhtml |
| 2021/01/11 | Hebei | Shijiazhuang | 39 | wsjkw.hebei.gov.cn/syyctplj/375429.jhtml |
| 2021/01/11 | Hebei | Langfang | 1 | wsjkw.hebei.gov.cn/syyctplj/375428.jhtml |
| 2021/01/12 | Hebei | Shijiazhuang | 84 | wsjkw.hebei.gov.cn/syyctplj/375478.jhtml |
| 2021/01/12 | Hebei | Xingtai | 6 | wsjkw.hebei.gov.cn/syyctplj/375477.jhtml |
| 2021/01/13 | Hebei | Shijiazhuang | 75 | wsjkw.hebei.gov.cn/syyctplj/375501.jhtml |
| 2021/01/13 | Hebei | Xingtai | 6 | wsjkw.hebei.gov.cn/syyctplj/375500.jhtml |
| 2021/01/14 | Hebei | Shijiazhuang | 84 | wsjkw.hebei.gov.cn/syyctplj/375548.jhtml |
| 2021/01/14 | Hebei | Xingtai | 6 | wsjkw.hebei.gov.cn/syyctplj/375547.jhtml |
| 2021/01/15 | Hebei | Shijiazhuang | 83 | wsjkw.hebei.gov.cn/syyctplj/375547.jhtml |
| 2021/01/15 | Hebei | Xingtai | 7 | wsjkw.hebei.gov.cn/syyctplj/375582.jhtml |
| 2021/01/16 | Hebei | Shijiazhuang | 65 | wsjkw.hebei.gov.cn/syyctplj/375582.jhtml |
| 2021/01/16 | Hebei | Xingtai | 7 | wsjkw.hebei.gov.cn/syyctplj/375618.jhtml |
| 2021/01/17 | Hebei | Shijiazhuang | 52 | wsjkw.hebei.gov.cn/syyctplj/375644.jhtml |
| 2021/01/17 | Hebei | Xingtai | 2 | wsjkw.hebei.gov.cn/syyctplj/375643.jhtml |
| 2021/01/18 | Hebei | Shijiazhuang | 35 | wsjkw.hebei.gov.cn/syyctplj/375680.jhtml |
| 2021/01/19 | Hebei | Shijiazhuang | 13 | wsjkw.hebei.gov.cn/syyctplj/375719.jhtml |
| 2021/01/19 | Hebei | Xingtai | 6 | wsjkw.hebei.gov.cn/syyctplj/375718.jhtml |
| 2021/01/20 | Hebei | Shijiazhuang | 20 | wsjkw.hebei.gov.cn/syyctplj/375757.jhtml |

| | | | | |
|---|---|---|---|---|
| 2021/01/21 | Hebei | Shijiazhuang | 15 | wsjkw.hebei.gov.cn/syyctplj/375796.jhtml |
| 2021/01/21 | Hebei | Xingtai | 3 | wsjkw.hebei.gov.cn/syyctplj/375795.jhtml |
| 2021/01/22 | Hebei | Shijiazhuang | 11 | wsjkw.hebei.gov.cn/syyctplj/375795.jhtml |
| 2021/01/22 | Hebei | Xingtai | 4 | wsjkw.hebei.gov.cn/syyctplj/375834.jhtml |
| 2021/01/23 | Hebei | Shijiazhuang | 17 | wsjkw.hebei.gov.cn/syyctplj/375834.jhtml |
| 2021/01/23 | Hebei | Xingtai | 2 | wsjkw.hebei.gov.cn/syyctplj/375834.jhtml |
| 2021/01/24 | Hebei | Shijiazhuang | 7 | wsjkw.hebei.gov.cn/syyctplj/375834.jhtml |
| 2021/01/24 | Hebei | Xingtai | 4 | wsjkw.hebei.gov.cn/syyctplj/375834.jhtml |
| 2021/01/25 | Hebei | Shijiazhuang | 5 | wsjkw.hebei.gov.cn/syyctplj/375834.jhtml |
| 2021/01/26 | Hebei | Shijiazhuang | 5 | wsjkw.hebei.gov.cn/syyctplj/375834.jhtml |
| 2021/01/26 | Hebei | Dingzhou | 1 | wsjkw.hebei.gov.cn/syyctplj/375834.jhtml |
| 2021/01/26 | Hebei | Xingtai | 1 | wsjkw.hebei.gov.cn/syyctplj/375834.jhtml |
| 2021/01/27 | Hebei | Shijiazhuang | 2 | wsjkw.hebei.gov.cn/syyctplj/375834.jhtml |
| 2021/01/27 | Hebei | Xingtai | 1 | wsjkw.hebei.gov.cn/syyctplj/375834.jhtml |
| 2021/01/28 | Hebei | Shijiazhuang | 1 | wsjkw.hebei.gov.cn/syyctplj/375834.jhtml |
| 2021/01/29 | Hebei | Shijiazhuang | 1 | wsjkw.hebei.gov.cn/syyctplj/375834.jhtml |
| 2021/01/30 | Hebei | Shijiazhuang | 1 | wsjkw.hebei.gov.cn/syyctplj/376102.jhtml |
| 2021/01/31 | Hebei | Shijiazhuang | 1 | wsjkw.hebei.gov.cn/syyctplj/376102.jhtml |
| 2021/02/02 | Hebei | Shijiazhuang | 1 | wsjkw.hebei.gov.cn/syyctplj/376194.jhtml |
| 2021/02/03 | Hebei | Shijiazhuang | 2 | wsjkw.hebei.gov.cn/syyctplj/376231.jhtml |
| | | | 941 | |

**Table A.1: List of data source of individual patient narratives published from 2nd January 2021 to 3rd February 2021 by Health Commission of Hebei Province**

### B. Examples of data extraction from four types of data

We provided one example for each of the four types of data to show extraction of relevant transmission timeline (in MM/DD/YYYY format) and covariates.

| Type | Patient narratives | Data extracted |
|---|---|---|
| I | Confirmed case 6: Female, 55 years old, currently living in Xiaoguozhuang Village, Gaocheng District. From December 20th to December 26th, 2020, the patient stayed at home and did not go out of the town. On December 27th, the patient walked to the village to participate in some activity, stayed there for about one hour, and then returned home by foot. On the morning of December 28th, she rode an electric bike to a hotel near North Airport Road to attend a wedding, but did not go out in the afternoon. The patient stayed at home and did not go out of the town from December 29th to 31st. After the supper on January 1st, 2021, due to headache and cough, she rode an electric bike to the village clinic to get some medicine from the doctor. Nucleic acid test was done on January 2nd. And she started home-isolation on January 2nd. The nucleic acid test result showed positive on January 3rd. On the same day, she was transferred to the Fifth Hospital of Shijiazhuang City by a negative pressure ambulance. She was diagnosed as a confirmed case on January 4th with grade of "moderate".<br><br>webpage of the original text in Chinese:<br>http://wsjkw.hebei.gov.cn/syyctplj/375192.jhtml | $W_0$= NA<br>$W_1$= NA<br>$T_0$=12/28/2020<br>$T_1$=01/01/2021<br>$T_2$=01/02/2021<br><br>Sex= female<br>Age= 55<br>Living with a case= no<br>Symptomatic= yes<br># of false negative= 0 |
| II | Confirmed case 14: Female, 24 years old, currently lives in Area A, District 4, Zhongmei Bridge community, Yuhua District. At 10 am on December 28th, 2020, she drove to Oujing Ecological Park, North Airport Road, Zhengding County to attend a wedding banquet. After that, she went to the hometown of Xiaoguozhuang Village, Zengcun Town, Gaocheng District, and then drove back home at 4 pm. From December 29th to January 31th, she rode an electric bicycle to the kindergarten in Area A of Anyuan Community to pick up and drop off her child. From January 1st to 2nd, 2021, except for some walks in the community, she did not visit any other public places. On January 3rd, she drove to the first affiliated hospital of Hebei Medical University of Hebei Province to take a nucleic acid test. The result was negative. On January 4th, the nucleic acid test showed positive. Then, she was transferred to the Fifth Hospital of Shijiazhuang City by the negative pressure ambulance on the same day. On January 9th, she was transferred to Hebei Chest Hospital by the negative pressure ambulance. She was diagnosed as a confirmed case on January 10th. | $W_0$= NA<br>$W_1$= NA<br>$T_0$=12/28/2020<br>$T_1$= censored<br>$T_2$=01/04/2021<br><br>Sex= female<br>Age= 24<br>Living with a case= no<br>Symptomatic= no<br># of false negative= 1 |

| | | |
|---|---|---|
| | webpage of the original text in Chinese: http://wsjkw.hebei.gov.cn/syyctplj/375391.jhtml | |
| III | Confirmed case 18: Male, 51 years old, lives in Nanqiaozhai Village, Zengcun Town, Gaocheng District. He was self-employed for house decoration work in Changping, Beijing. He returned home from Beijing on December 29th, 2020. He did not go out of the village from December 30th, 2020 to January 2nd, 2021. On January 3rd, he had fever and took a nucleic acid test on the same day. The result showed positive on January 4th. On January 5th, he was transferred to the Fifth Hospital of Shijiazhuang City by a negative pressure ambulance. He was diagnosed as confirmed case on the same day.<br><br>webpage of the original text in Chinese: http://wsjkw.hebei.gov.cn/syyctplj/375221.jhtml | $W_0$=12/29/2020<br>$W_1$=01/02/2021<br>$T_0$=NA<br>$T_1$=01/03/2021<br>$T_2$=01/05/2021<br><br>Sex= male<br>Age= 51<br>Living with a case= no<br>Symptomatic= no<br># of false negative= 0 |
| IV | Confirmed case 7: female, 49 years old, currently lives in Ruida Rose Garden community, Changshou street, Xinle City, the mother of the 25th confirmed case on January 17th. From January 1st to January 4th, 2021, she rode an electric bike to work at the cloth shop in Xianyu market, Xinle City every day at 9am, returned home at 4pm. She was transferred to a designated isolation place in Xinle City for medical observation on January 5th. The nucleic acid tests on January 5th, 7th, 9th, and 13th were all negative. On January 15th, the nucleic acid test showed positive. On January 16th, she was transferred by a negative pressure ambulance to Shijiazhuang People's Hospital of Jianhua District and she was diagnosed as asymptomatic case. On January 19th, she was diagnosed as confirmed case.<br><br>webpage of the original text in Chinese: http:// wsjkw.hebei.gov.cn/syyctplj/375719.jhtml | $W_0$= NA<br>$W_1$=01/04/2021<br>$T_0$=NA<br>$T_1$= censored<br>$T_2$=01/15/2021<br><br>Sex= female<br>Age=49<br>Living with a case= yes<br>Symptomatic= no<br># of false negative= 4 |

### C. Summary of 20 publications about median incubation period

| Article | Coverage period (MM/DD/YYYY) | Data source region | N | Median | Distribution model (analysis software) | Study type |
|---|---|---|---|---|---|---|
| Backer et al. (2020)[1] | 01/20/2020-01/28/2020 | China | 88 | 6·4 | Weibull | meta-analysis |
| Bui et al. (2020)[2] | 01/23/2020-04/13/2020 | Vietnam | 19 | 5·6 | Weibull | independent study |
| Cheng et al. (2020)[3] | Before 03/18/2020 | Taiwan | 100 | 4·1 | NA | independent study |
| Du et al. (2020)[4] | 01/08/2020-02/05/2021 | China | 109 | 5·1 | Gamma | independent study |
| Deng et al. (2020)[5] | 01/19/2020-01/23/2020 | China | 1211 | 8·5 | Gamma | independent study |
| Guan et al. (2020)[6] | 01/12/2019-01/29/2020 | China | 1099 | 4 | NA | independent study |
| Kong et al. (2020)[7] | 01/15/2020-01/22/2020 | China | 136 | 8·3 | NA (CumFreq) | independent study |
| Lauer et al. (2020)[8] | 01/04/2020-02/24/2020 | China, except Hubei province | 181 | 5·1 | Lognormal | independent study |
| Lee et al. (2020)[9] | 02/20/2020-03/03/2020 | Korea | 47 | 3 | Lognormal | independent study |
| Linton et al. (2020)[10] | 01/01/2020-01/31/2021 | China, except Wuhan | 158 | 5·6 | Lognormal | independent study |
| McAloon et al. (2020)[11] | 12/01/2019-04/08/2020 | Worldwide | NA | 5·1 | Lognormal | meta-analysis |
| Nie et al. (2020)[12] | 01/19/2020-02/08/2020 | China, except Hubei province | 7015 | 5 | NA | independent study |
| Patrikar et al. (2020)[13] | 02/01/2020-04/19/2020 | India | 268 | 6.93 | Normal | independent study |
| Qian et al. (2020)[14] | 01/20/2020-02/11/2020 | Zhejiang, China | 91 | 6 | NA | independent study |
| Qin et al. (2020)[15] | Before 02/15/2020 | China, except Hubei province | 1084 | 7·76 | Weibull | independent study |
| Sun et al. (2020)[16] | 01/20/2020-0 | Beijing, | 55 | 7·5 | NA | independent |

| | | | | | | |
|---|---|---|---|---|---|---|
| | 2/15/2020 | China | | | | study |
| Tan et al. (2020)[17] | 01/23/2020-04/02/2020 | Singapore | 164 | 5 | NA | independent study |
| Viego et al. (2020)[18] | 03/20/2020-05/08/2020 | Argentina | 18 | 6·1 | Lognormal | independent study |
| Xiao et al. (2020)[19] | Before 02/21/2020 | China, except Hubei, Qinghai, Xizang Province | 2555 | 8 | Weibull | independent study |
| Yang et al. (2020)[20] | 01/20/2020-04/01/2020 | Shiye, Hubei province | 178 | 5·4 | Weibull (coarseDataTools) | independent study |

**Table C.1: Summary of 20 publications about median incubation period used in plotting Figure 4 and meta-analysis**

### D. Details of the meta-analysis about median incubation period

A simple meta-analysis based on seven published articles was conducted to obtain a pooled estimate of the median of incubation period (IP) during the first SARS-CoV-2 pandemic wave (from December 2019 to April 2020) in China. The process was outlined by the flow chart in Figure D.1.

The R package "metamedian" developed by McGrath[21] was used to analyze the data. The detail of the source information along with the forest plot was shown in Figure D.2. The estimate of the overall median IP was found to be 5·6 days (95% CI 5·1–8·3).

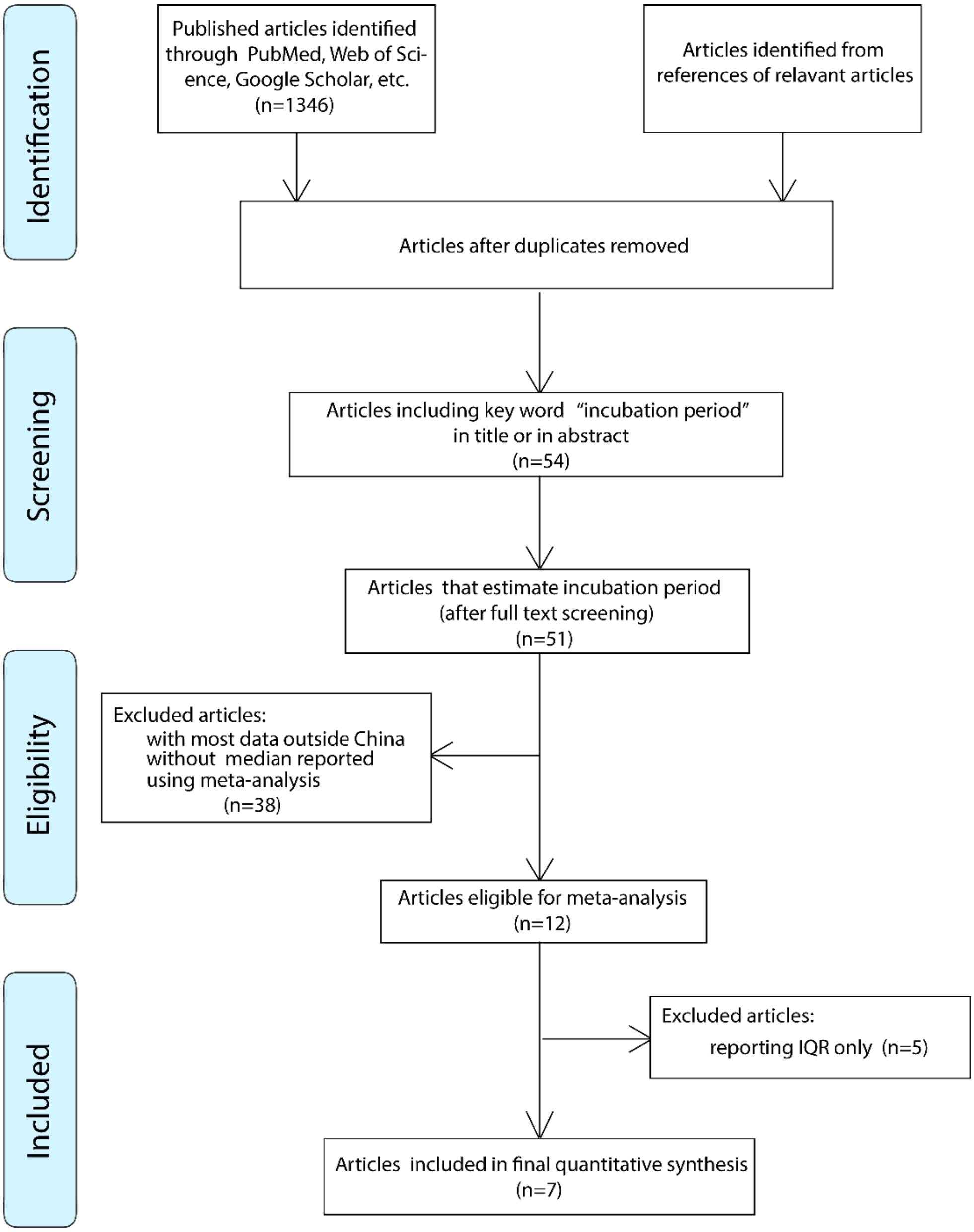

**Figure D.1: Flow chart of processing of source information for the meta-analysis**

| Study | Sample Size | Median | 95% CI |
|---|---|---|---|
| Deng et al. (2020) | 1211 | 8.5 | [7.22,9.15] |
| Kong et al. (2020) | 136 | 8.3 | [7.6,9.0] |
| Lauer et al. (2020) | 181 | 5.1 | [4.5,5.8] |
| Linton et al. (2020) | 158 | 5.6 | [4.9,6.4] |
| McAloon et al. (2020) | - | 5.1 | [4.5,5.8] |
| Qin et al. (2020) | 1084 | 7.76 | [7.02,8.53] |
| Yang et al. (2020) | 178 | 5.4 | [4.8,6.0] |
| **Summary** | | **5.6** | **[5.1,8.3]** |

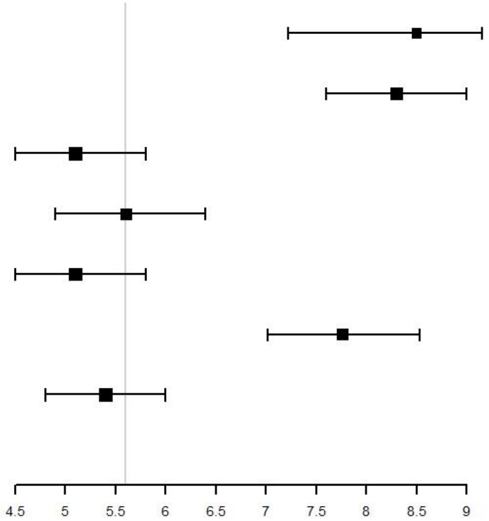

**Figure D.2: Source information used in the meta-analysis and the resulting forest plot for the overall estimate of the median incubation period**

### E. Details of analyzing covariate effects

We applied the Cox model to test the covariate effects of sex, age and living with a case with respect to IP. The estimated coefficients of covariates and associated p-values were obtained in Table E.1 based on different aggregated types of data. All p-values were greater than 0 05, indicating no significant effect of the considered covariates.

| aggregated types | sex | age | living with a case |
|---|---|---|---|
| I + II | 0.016 (0.10) | -0.44 (0.29) | -0.42 (0.32) |
| I + II + III | 0.016 (0.10) | -0.50 (0.22) | -0.52 (0.22) |
| I + II + III + IV | 0.015 (0.11) | -0.49 (0.11) | -0.25 (0.56) |

**Table E.1: Estimated coefficients of covariates and associated p-values in parentheses obtained by the Cox model for the different aggregated types of data**

We used the traditional log-rank test to test the equality of the distributions of IP between two age groups. Figure E.1 shows Kaplan-Meier estimates of the probability of being asymptomatic of the two age groups based on different aggregated types of data. The p-values of the test were shown in the second column of Table E.2. In the presence of possible crossing survival curves, we also reported the p-values in the third column of Table E.2 of the restricted mean survival time (RMST) based test[22] between the two groups. Again, none of the results showed significant difference between the two groups.

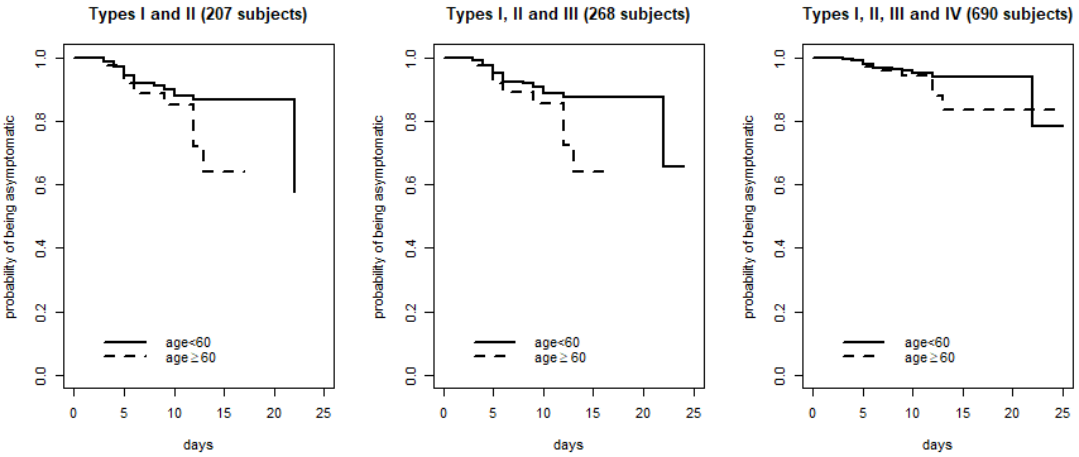

**Figure E.1: Kaplan-Meier estimates of probability of being asymptomatic of two age groups based different aggregated types of data**

| aggregated types | Log-rank test | RMST-based test |
|---|---|---|
| I + II | 0.09 | 0.16 |
| I + II + III | 0.07 | 0.13 |
| I + II + III + IV | 0.20 | 0.29 |

**Table E.2: P-values of the log-rank test (for equality of the distributions of IP) and p-values of the RMST-based test (for equality of RMSTs of IP)**
